# Differences in outcomes following an intensive upper-limb rehabilitation programme for patients with common CNS-acting drug prescriptions

**DOI:** 10.1101/2020.10.21.20215038

**Authors:** Ainslie Johnstone, Fran Brander, Kate Kelly, Sven Bestmann, Nick Ward

## Abstract

Difficulty using the upper-limb is a major barrier to independence for many patients post-stroke or brain injury. High dose rehabilitation can result in clinically significant improvements in function even years after the incident, however there is still high variability in patient responsiveness to such interventions that cannot be explained by age, sex or time since stroke.

This retrospective study investigated whether patients prescribed certain classes of CNS-acting drugs - GABA agonists, antiepileptics and antidepressants-differed in their outcomes on the 3 week intensive Queen Square Upper-Limb (QSUL) programme.

For 277 stroke or brain injury patients (167 male, median age 52 years (IQR 21), median time since incident 20 months (IQR 26)) upper-limb impairment and activity was assessed at admission to the programme and at 6 months post-discharge, using the upper limb component of the Fugl-Meyer (FM), Action Research Arm Test (ARAT), and Chedoke Arm and Hand Activity Inventory (CAHAI). Drug prescriptions were obtained from primary care physicians at referral. Specification curve analysis (SCA) was used to protect against selective reporting results and add robustness to the conclusions of this retrospective study.

Patients with GABA agonist prescriptions had significantly worse upper-limb scores at admission but no evidence for a significant difference in programme-induced improvements was found. Additionally, no evidence of significant differences in patients with or without antiepileptic drug prescriptions on either admission to, or improvement on, the programme was found in this study. Whereas, though no evidence was found for differences in admission scores, patients with antidepressant prescriptions experienced reduced improvement in upper-limb function, even when accounting for anxiety and depression scores.

These results demonstrate that, when prescribed typically, there was no evidence that patients prescribed GABA agonists performed worse on this high-intensity rehabilitation programme. Patients prescribed antidepressants, however, performed poorer than expected on the QSUL rehabilitation programme. While the reasons for these differences are unclear, identifying these patients prior to admission may allow for better accommodation of differences in their rehabilitation needs.

## Introduction

Stroke is the most common cause of long-term neurological disability worldwide (1). Currently, half of all people who survive a stroke are left disabled, with a third relying on others to assist with activities of daily living (2). A major contributor to ongoing physical disability is persistent difficulty in using the upper-limb (3). For many years it was believed that spontaneous upper-limb recovery occurred in the first 3 months following a stroke, with only small rehabilitation-induced improvements happening after this period (4). However, recent studies have demonstrated that with specific, high-dose training chronic patients can experience clinically significant improvements in upper-limb function (5–7). Yet despite these positive results, there is a degree of variability in patient outcomes that cannot be explained by impairment at admission or other patient characteristics (7). Identifying factors influencing this variability is therefore of high priority if similar high-intensity interventions are to be effectively developed.

There is an increasing wealth of literature, in both animals and humans, indicating that certain commonly used prescription drugs influence motor recovery following a brain lesion. Experimental findings from humans (8–12) indicate that selective serotonin reuptake inhibitors (SSRIs) may boost practice-dependent motor improvements, while animal experiments (13,14) and retrospective human studies (15,16) indicate activation at GABA receptors is detrimental to motor recovery. Though carefully matched placebo-controlled studies are the gold-standard for identifying the true effects of a given drug on motor recovery, these trials are costly and practically difficult. They must combine chronic drug administration with specific high-dose motor training (17).

Retrospective analysis that examines the relationship between drug prescriptions and patients’ response to rehabilitation programmes can provide a solution to some of these issues. In a naturalistic setting, prescriptions of common drugs come hand-in-hand with the co-morbidities they are aiming to treat, such as depression, epilepsy or spasticity. These issues may themselves impact on recovery, or interact with effects of the drug, making it difficult to draw conclusions about specific drug effects. However, using drug prescriptions to identify patients who systematically respond better or worse to a given intervention is the first step to singling out the causes of these disparities, and eventually leveraging these findings to improve interventions for all.

Another potential issue surrounding retrospective analysis of existing datasets is that, without pre-registration, researchers can be biased to make arbitrary analysis decisions motivated by results, rather than theory. A novel method, known as specification curve analysis (SCA), has been developed to tackle this problem (18). Using SCA, all reasonable variations of a possible analytical test assessing each hypothesis are run. Rather than examining the results of individual tests, the results across all tests are interpreted together to make a decision about whether to reject the null hypothesis (18).

## Aims

This retrospective study used SCA analysis to examine whether patients with prescriptions for certain classes of common drugs acting on the central nervous system (CNS) (i) differed in their level of upper-limb impairment on admission to a high-dose Queen Square Upper-Limb (QSUL) rehabilitation programme and (ii) differed their response to the programme. The drug categories examined were GABA agonists, antiepileptics acting on sodium or calcium channels, and antidepressants.

## Methods

### Patient Data

Patients were referred to the QSUL programme by primary care physicians. The inclusion criteria for admission to the program was/is broad, focussing on whether patients were likely to achieve their goals for their upper-limb. There were no restrictions on time since stroke/injury or other demographic factors, but for patients who experienced any of the following high intensity rehabilitation was considered unlikely to be beneficial: i) no active movement in shoulder flexion/forward reach or hand opening/finger extension; (ii) a painful shoulder limiting an active forward reach (mostly due to adhesive capsulitis); (iii) severe spasticity or non-neural loss of range and (iv) unstable medical conditions. For more information regarding patient admission see Ward et al., 2019.

Between April 2014 and March 2020, a total of 439 first-time patients had been admitted to the 3-week programme. Of these, 321 patients had completed the 6 week and 6 month follow-up. There were several reasons that patients were not available for follow-up: some could not be contacted, considered it too far to travel, or suffered intercurrent illnesses; a large number were due for follow-up after the UK COVID-19 lockdown in March 2020. A further 15 patients were excluded as they did not have mood and/or fatigue measures recorded, and a final 29 patients were excluded as prescription drug information was not supplied at referral. This left a total of 277 patients for whom full data sets were available. A break-down of demographics of the included 277 patients and the excluded 162 are provided in table 1.

**Table 1:** Admission information for included and excluded patients.

|  | Included patients,<br>n=277 | Excluded patients,<br>n=161 | Statistical comparison |
| --- | --- | --- | --- |
| Age in years, median (IQR, range) | 52 (21, 16-79) | 54 (19, 16-84) | W(161, 277)=20184, p=0.098 |
| Gender, male | 167 | 101 | $\chi^2(1)=0.164$, p=0.686 |
| Time since incident in months,<br>median (IQR, range) | 20 (26, 2-340) | 18 (21 2-409) | W(161, 277)=23444, p=0.370 |
| Lesion type: Hemorrhagic | 76 (27%) | 41 (25%) | $\chi^2(2)=5.84$, p=0.054 |
| Ischemic | 172 (62%) | 90 (56%) |  |
| Other/unknown | 29 (10%) | 30 (19%) |  |
| Affected limb, right | 140 | 86 | $\chi^2(1)=0.518$, p=0.472 |
| Dominant limb affected | 143 | 88 | $\chi^2(1)=0.261$, p=0.607 |
| Admission Barthel Index, median<br>(IQR) | 19 (2) | 18 (2) | W(161, 277)=22525, p=0.240 |
| HADS score, median (IQR) | 12 (8) | 14 (12) | W(161, 277)=17226, **p=0.012** |
| NFI score, median (IQR) | 35 (15) | 40 (14) | W(161, 277)=15489, **p<0.001** |
| Drug prescriptions: GABA agonists | 49 (18%) | 20/117 (17%) | $\chi^2(2)=1.051$, p=0.591 |
| Antiepileptics | 81 (29%) | 46/117 (39%) |  |
| Antidepressants | 56 (20%) | 30/117 (26%) |  |

*IQR- Interquartile range; HADS- Hospital anxiety and depression scale; NFI -Neurological Fatigue Index.*

### Upper-limb Measures

Function of the affected upper-limb was assessed on admission, discharge, 6 weeks and 6 months post-discharge using the following measures: Fugl-Meyer upper-limb (FM), Action Research Arm Test (ARAT), and the Chedoke Arm and Hand Activity Inventory (CAHAI). The FM is a stroke-specific, performance-based impairment index. Here a modified version was used-excluding coordination and reflexes-which specifically focussed on motor synergies and joint function. This had a maximum score of 54 and the minimum clinically important difference (MCID) has been reported as 5.25 points (19). The ARAT assesses patients’ ability to handle objects of differing size, weight and shape. It has a maximum score of 57 and a MCID of 5.7 points (20). Finally, the CAHAI focuses on how the arm and hand are incorporated into bilateral activities of daily living. The maximum score is 91 and though no MCID has been reported the minimum detectable change has been reported as 6.2 points (21).

### Additional Demographic or Subjective Measures

At admission two subjective measures, the Hospital Anxiety and Depression Scale (HADS) and the Neurological Fatigue Index (NFI), scored out of 42 and 69 respectively, were administered. Other demographic information, e.g. age and sex, and neurological information, e.g. time since stroke/injury (at admission) and whether their dominant arm was affected, was also recorded.

Primary care physicians supplied each patient’s prescribed drugs at the time of referral. Drugs acting on the CNS were grouped into three categories: GABA agonists, antiepileptics (acting on sodium or calcium channels), and antidepressants. Patients were coded as ‘on’ a category if they prescribed one (or more) of the drugs within the category. Dose or prescription directions were not recorded. The specific drugs included in each category were: GABA agonists (n=49) – baclofen (n=41), clonazepam (n=3), diazepam (n=4), clobazam (n=2), and sodium valproate (n=3); antiepileptics (n=81) – topiramate (n=1), zonisamide (n=2), lamotrigine (n=13), lacosamide (n=4), (ox)carbazepine (n=2), phenytoin (n=3), levetiracetam (n=33), pregabalin (n=16) and gabapentin (n=21); antidepressants (n=56) - fluoxetine (n=9), citalopram (n=20), escitalopram (n=1), sertraline (n=10), paroxetine (n=2), duloxetine (n=2), venlafaxine (n=1), mirtazapine (n=9) and amitriptyline (n=9). While there are other centrally acting drug categories that would have been of interest, they were not prescribed in sufficient numbers to make analysis viable (e.g. neuroleptics n=3, cholinergic drugs n=0, dopaminergic drugs n=3, centrally acting hypertensives n=1)

## Analysis

All analyses were performed using R (RStudio version 1.1.456). Though this study had the clear objective of testing whether patients prescribed certain classes of CNS-acting drug prescriptions differed in motor outcomes following the QSUL programme, as a retrospective analysis of existing data, pre-registration was not a convincing solution to eliminating bias in subjective analysis decisions. Increasingly, specification curve analyses (SCA) are being used to circumvent this problem for hypothesis testing on medium-to-large data sets (18,22–24). SCA is a tool for mapping out a relationship of interest across all potential, defensible, hypothesis tests examining this relationship. Conclusions are drawn from the sum total of the results across all of the analyses rather than focussing on the results of only one test. While this method could be criticised for lumping together multiple different hypotheses in this case our overarching theoretical hypothesis, that there is a relationship between drug prescriptions and motor outcomes-a concept which is assessed by all three upper-limb measures-makes the SCA well suited.

SCAs were run on a variety of linear regression models examining whether patients in certain drug prescription groups - GABA agonists, antiepileptics, and antidepressants - differed on (i) admission motor function and/or (ii) recovery/outcome at the 6 month timepoint. To assess the differences across the drug groups, the regression coefficient (i.e. the magnitude of the relationship between prescription group and the admission score) and the p-value (i.e. whether this relationship was statistically significant) were extracted from each of the linear models and fed into the SCA. The code is available here [https://github.com/ainsliej/SCA-QSUL_Drugs].

### Identification of individual models for specification

For each of the three upper-limb measures-FM, ARAT and CAHAI-the association between the score at admission and the drug group was estimated using a linear regression model containing the prescription drug of interest and a variety of different covariates, grouped in pairs, which could be included or excluded from the analyses. These were: demographic information (i.e. age and sex); neurological incident information (i.e. time since incident and whether the dominant arm was primarily affected); subjective measures (i.e. HADS and NFI); and prescription of the other two drug groups. Inclusion or exclusion of outlying patients was also varied, where outlying patients were defined as having a recovery score (T_admission_ to T_6month_) that was outside 2.5*the interquartile range (IQR) from the median. This created a total of 96 different models, all assessing whether patients with prescriptions of the drugs of interest differed in upper-limb function at admission. To allow easier comparison between the different upper-limb measures, each of which has a different scale, all measures were converted to a proportion of the maximum score (T_x_/T_Max_).

To assess the association between drug prescriptions and improvement, all three upper-limb measures were again examined, and the same set of covariates were either included or excluded. There are a variety of different ways improvement could be modelled: an outcome model, examining the final T_6month_ score from the T_admission_ score; an absolute recovery model, examining the change in score from T_admission_ to T_6month_; or a relative recovery model, examining the amount of recovery achieved relative to the amount possible ((T_6month_ - T_admission_)/(Max Score - T_admission_)). This creates a total of 288 possible models all of which test the hypothesis that motor improvement following the QSUL differs by drug prescription status. Again, all outcome scores were proportions of the maximum possible score, and recovery scores were calculated using these proportions.

SCA models were also run to test whether patient’s HADS score was associated with improvement. The same models were run as for the drug prescription analysis, except all drugs were either included or excluded together, and NFI was included or excluded independent to HADS score.

### Hypothesis testing of SCA

In each SCA, a certain proportion of the models examined will report a relationship that reaches statistical significance (p<0.05). However, SCA aims to examine the evidence as a whole, summing across all the different individual models. In order to assess the statistical significance of the sum of evidence from a given SCA, a permutation method was used to generate the distribution of p-values, given the null hypothesis that the dependent variable (drug prescription) of interest has no relationship with the independent variable (admission/improvement score) (22). For each SCA, in 500 permutations, the independent variables were shuffled, while keeping the dependent variables and covariates un-shuffled. The total number of models with a significant relationship between the dependant and independent variable, for each permutation of the SCA was then extracted. A p-value for each SCA was calculated as the proportion of these permutations that had at least as many significant models as the original data.

## Results

### Differences between included and excluded participants

To assess whether there were any differences in the demographics of participants who were included in the analysis compared with those who were excluded, Mann-Whitney U and chi-square tests were performed, with full results reported in Table 1. Nominal variables were analysed using a non-parametric method as Shapiro-Wilks test indicated that all variables deviated from the normal distribution. Briefly, included participants tended to have lower HADS (W(161,277)=17226, p=0.012) and lower NFI (W(161,277)=15489, p<0.001) scores, but there was not sufficient evidence to reject the null hypothesis of no differences in any other measures. While these findings indicate that included participants were less depressed/anxious and had less fatigue, the median scores for both groups on HADS indicate mild depression/anxiety symptoms (25) and NFI scores were within a normal range (26).

### GABA agonist prescriptions had a significant negative relationship with admission scores, but not improvement

SCA of the admission scores revealed that patients who had a prescription of GABA agonists were significantly worse on admission to the QSUL (p<0.002). Of the 96 separate models run in the admission SCA, 84 reported a significant difference in scores between this drug category, and across all three of the different admission measures where patients with GABA agonist prescriptions had lower scores (see Figure 3A). The mean value of the regression coefficients (β) for significant results was -0.085, with a range of -0.115 to - 0.066. This equates to a mean of 8.5% (range 6.6 – 11.5%) reduction in admission scores in patients with a GABA agonist prescription relative to those without. Mean β across all models was -0.083 (range -0.115 to -0.062).

Using SCA to examine whether GABA agonist prescription related to degree of programme-related improvements in motor function did not generate sufficient evidence to reject the null hypothesis of no difference (p=0.266, 11/288 models significant, mean β= -0.026, range - 0.104 to 0.01; see Figure 2B).

**Figure 1:**
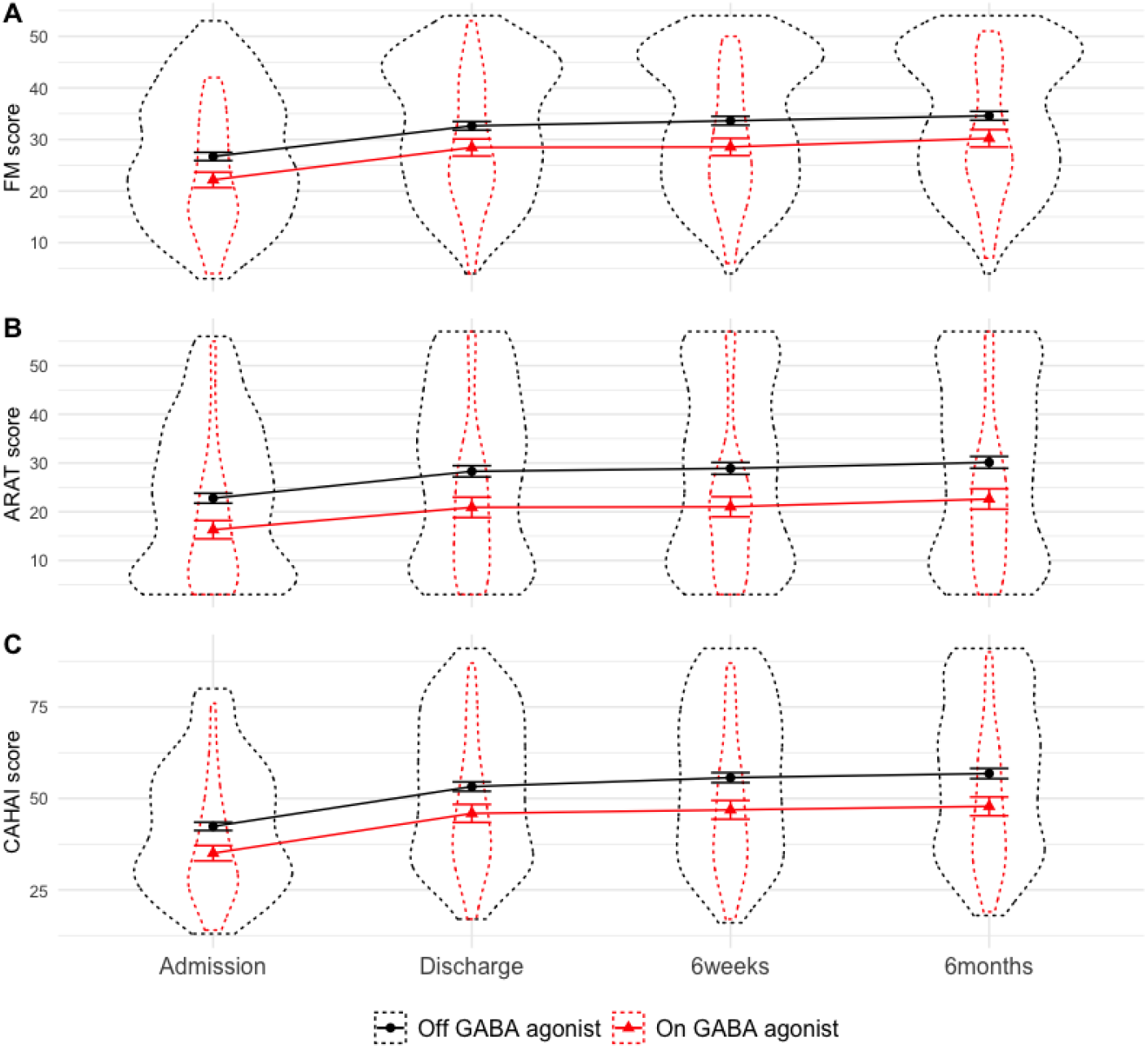
Measures of upper-limb function, across time split by GABA agonist prescription. Patients on GABA agonists had worse upper limb function at admission, but did not differ in degree of improvement during the programme. Dotted outline shows violin plot, solid lines show mean and standard error.

**Figure 2:**
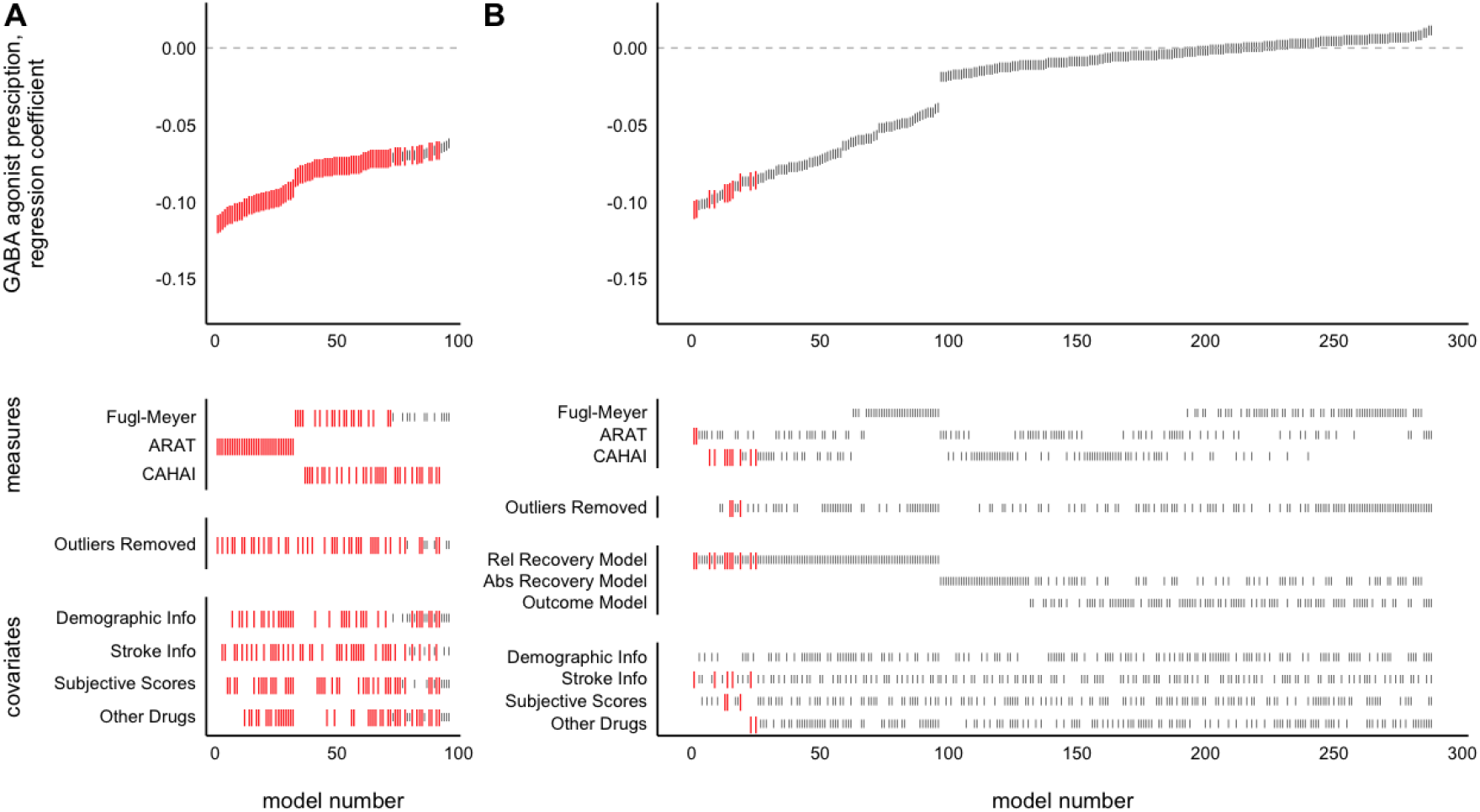
SCA examining relationship between GABA agonist prescription and measures of upper-limb function at admission (A) or improvement (B). Each model, sorted by the size of the GABA agonist prescription regression coefficient, is represented by a line in the top panel. Larger red lines represent a significant difference in scores across GABA agonist prescription groups. Lines in the lower panels indicate the contents of the model. Patients on GABA agonists had worse upper limb function at admission, but did not significantly differ in degree of improvement during the programme

**Figure 3:**
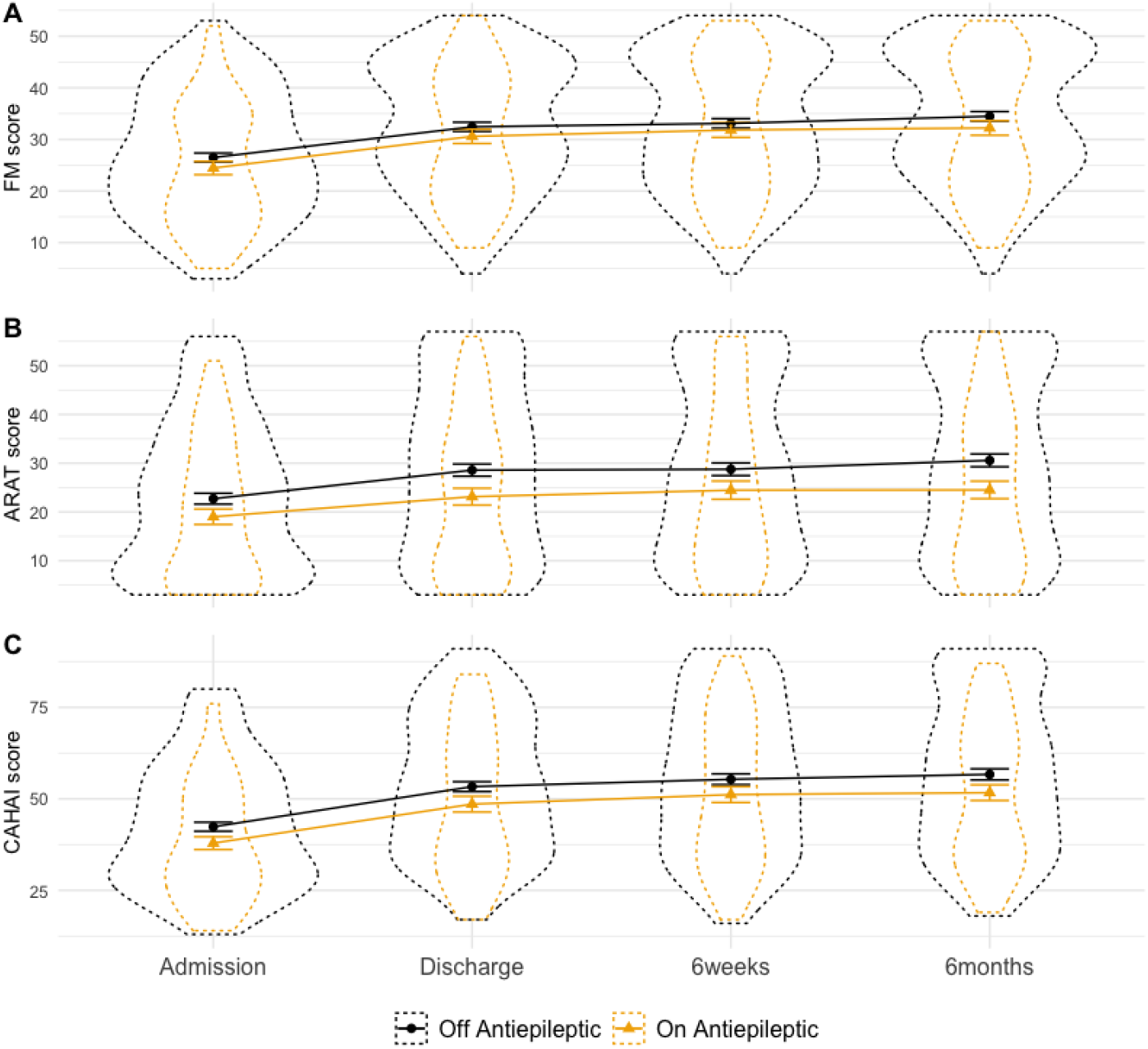
Measures of upper-limb function, across time split by antiepileptic prescription. Patients on and off antiepileptic drugs did not differ in admission or improvement scores. Dotted outline shows violin plot, solid lines show mean and standard error.

### No evidence of a significant relationship between antiepileptic prescriptions and admission scores or programme-related improvements

The results of the SCA revealed insufficient evidence to reject the null hypothesis of no relationship between antiepileptic prescription and admission scores (p=0.152, 2/96 models significant, mean β= -0.039, range -0.066 to -0.022). (see Figure 4A). However, SCA of antiepileptic prescription and improvements revealed a relationship approaching significance (p=0.052, 77/288 models significant, mean β= -0.032, range -0.159 to 0.006), driven by models examining ARAT scores.

**Figure 4:**
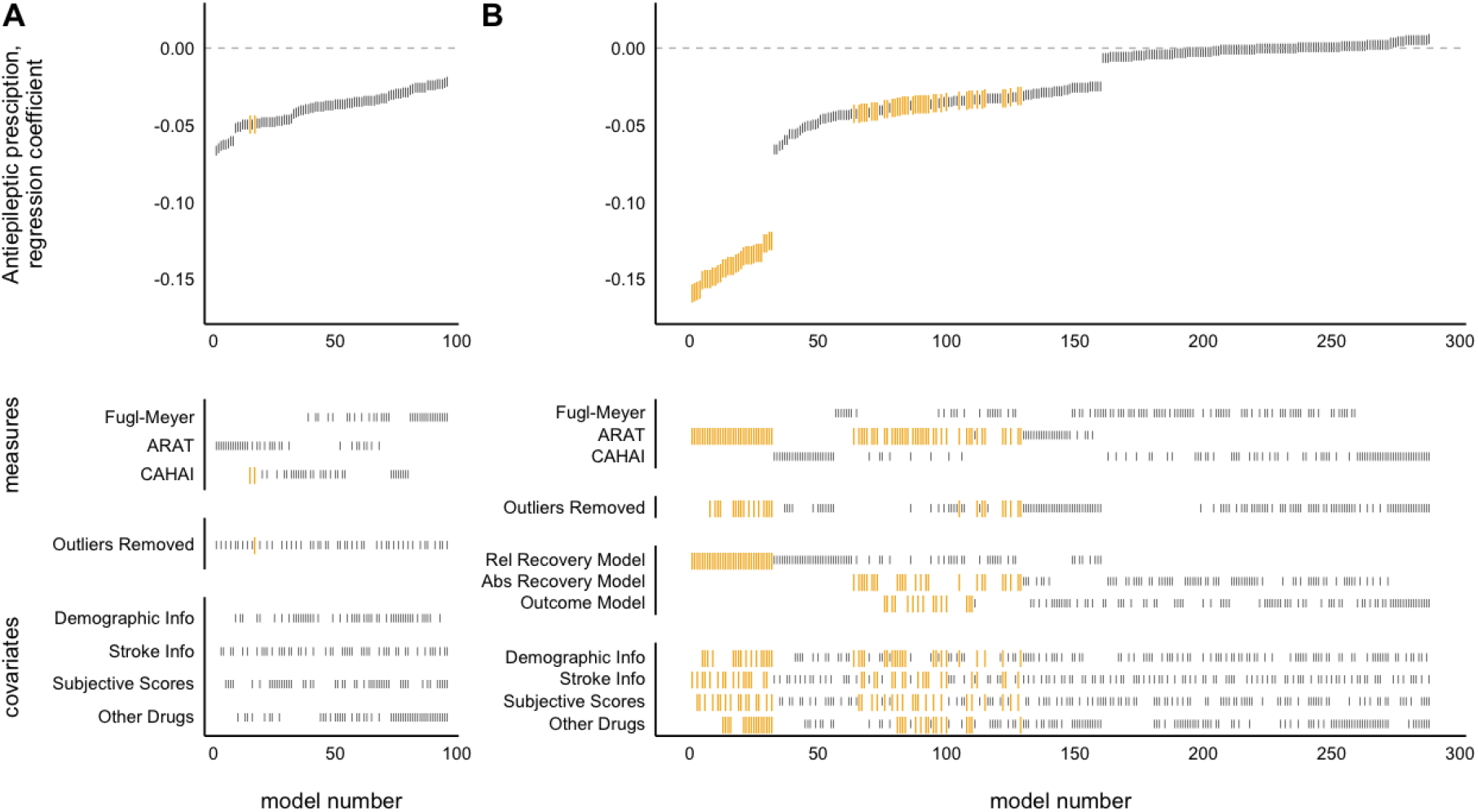
SCA examining relationship between antiepileptic prescription and measures of upper-limb function at admission (A) or improvement (B) Each model, sorted by the size of the antiepileptic prescription regression coefficient, is represented by a line in the top panel. Larger yellow lines represent a significant difference between scores in patients grouped by antiepileptic prescription. Lines in the lower panels indicate the contents of the model. Patients on and off antiepileptic drugs did not differ in admission or improvement scores.

### Antidepressant prescriptions had a significant negative relationship with improvement on QSUL

There was not sufficient evidence found using the SCA to reject the null hypothesis of no relationship between antidepressant prescription and admission scores (p=0.094, 13/92 models significant, mean β= -0.058, range -0.076 to -0.041). However, the SCA found evidence of a worsening of programme-related improvements in patients on antidepressants (p=0.016, 143/288 models significant, mean β= -0.047, range -0.127 to -0.010). Significant regression coefficients were found across all measures, though predominantly in FM and ARAT. The magnitude of regression coefficients was higher using the recovery model, but a similar number of significant results were found across all model types. Covariate inclusion did not appear to reliably dictate model significance or regression coefficient size.

### Patients with antidepressant prescriptions had higher HADS scores than those without

Although including subjective measures (i.e. HADS and NFI scores) did not systematically alter the significance or regression coefficient magnitude of the drug prescription relationship, we wanted to further examine the relationship between drug prescriptions and HADS score. Patients with antidepressant prescriptions had significantly higher depression/anxiety scores, as assessed by two-sample t-test of HADS scores, than those without (t(88)=2.76, p=0.007) (see Figure 6A). This was not however the case for GABA agonist (t(66)=1.46, p=0.148) or antiepileptic prescriptions (t(136)=1.01, p=0.312). NFI score also did not differ by antidepressant prescription (t(91)=0.80, p=0.425).

**Figure 5:**
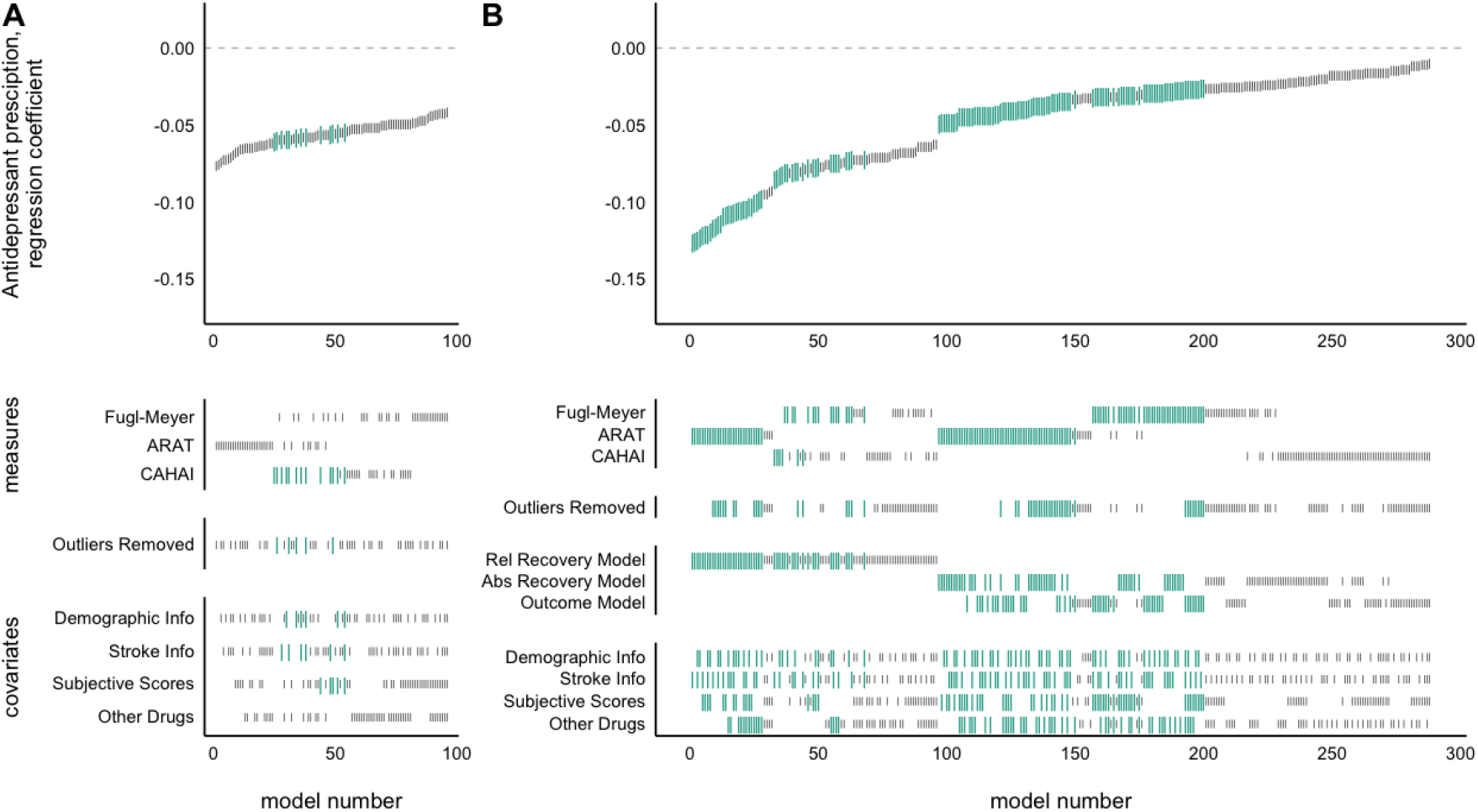
SCA examining relationship between antidepressant prescription and measures of upper-limb function at admission (A) or improvement (B) Each model, sorted by the size of the antidepressant prescription regression coefficient, is represented by a line in the top panel. Larger turquoise lines represent a significant difference between scores in patients grouped by antidepressant prescription. Lines in the lower panels indicate the contents of the model. Patients with antidepressant prescription did not differ in admission scores, but had lower programme-induced improvement scores.

**Figure 6:**
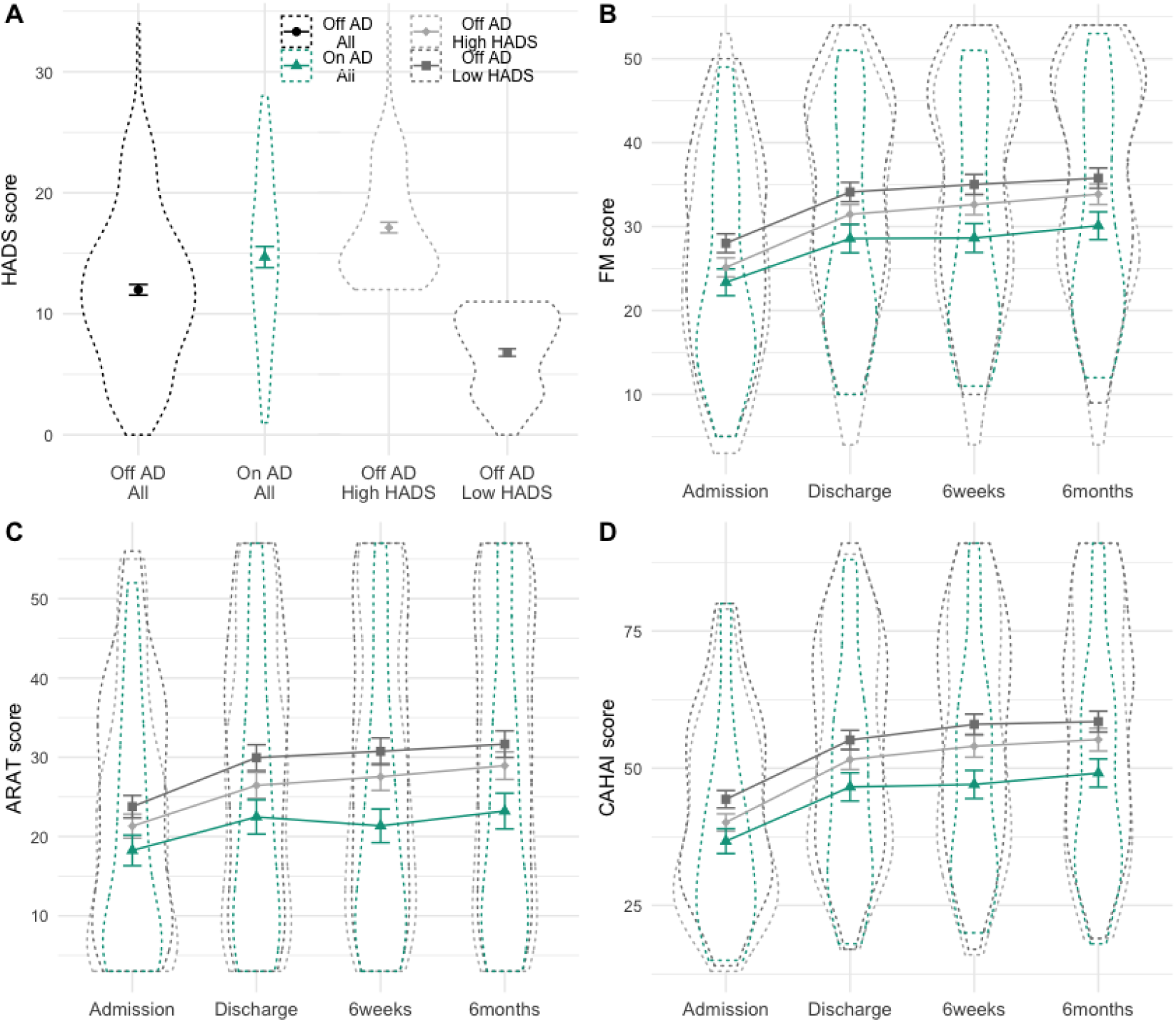
HADS score and upper-limb function scores split by antidepressant prescription. A, HADS scores for patients split by antidepressant prescription (black, turquoise), showing patients with antidepressant prescription have significant higher HADS score than those without. HADS scores for patients without antidepressant prescriptions, median split by HADS score, are also shown (light and dark grey). These groups have respectively higher and lower HADS scores than the group on antidepressants. Dotted outlines are

**Figure 7:**
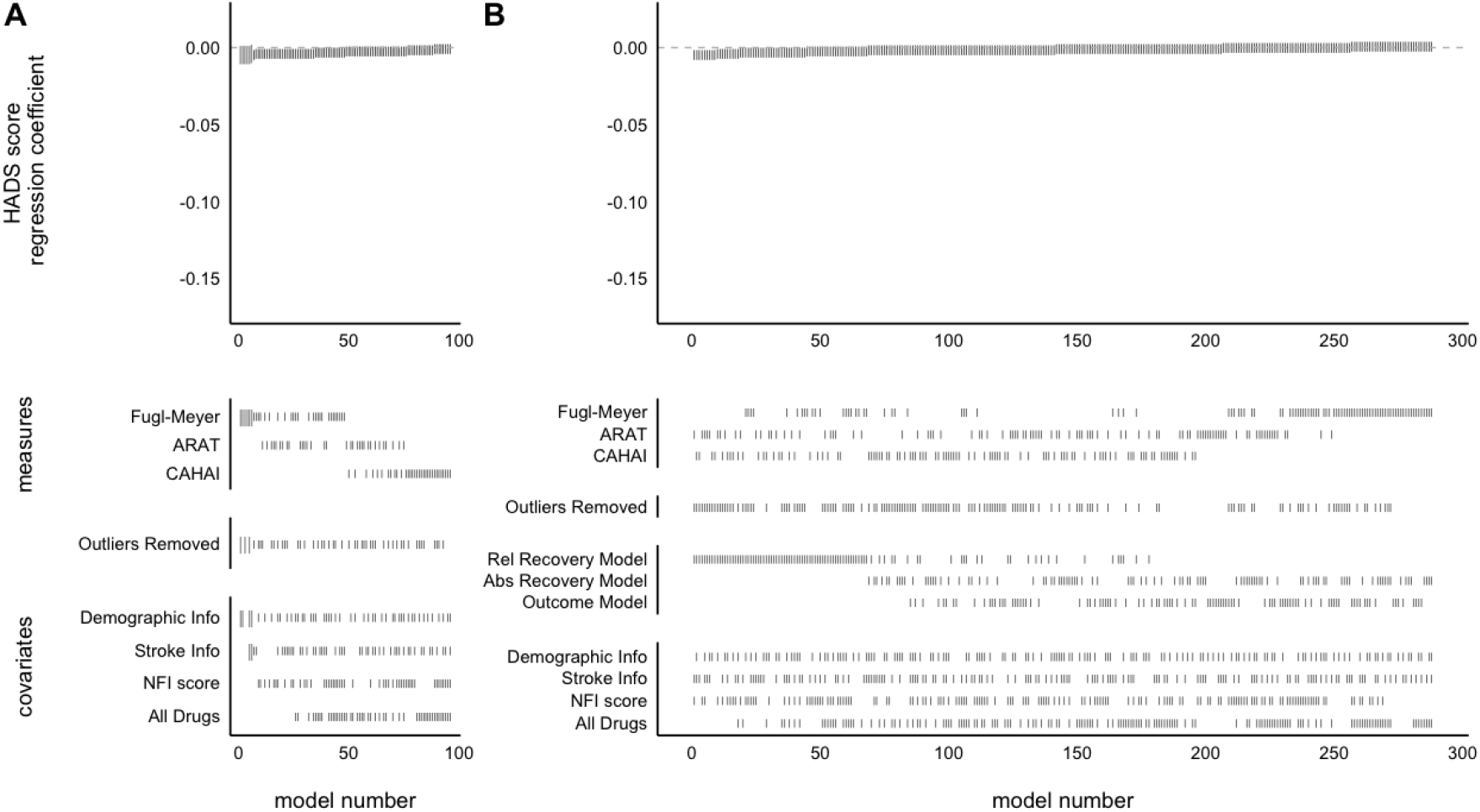
SCA of the relationship between HADS score and measures of upper-limb function at admission (A) or improvement (B) Each model, sorted by the size of the HADS score regression coefficient, is represented by a line in the top panel. Larger grey lines represent a significant relationship between HADS score and motor recovery/ outcome. Lines in the lower panels indicate the contents of the model. HADS score did not explain variance in baseline motor scores, or recovery/outcome scores.

To follow-up, a median split was performed on the HADS scores in patients without antidepressant prescription. These three groups (OnAD, OffAD-HighHADS, OffAD-LowHADS) had significantly different HADS scores (ANOVA: F(2,274)=142.3, p<0.001), and pairwise comparison showed that the AD+ group had significantly higher HADS score than the OffAD-LowHADS (Tukey HSD: diff=7.89, p<0.001) and significantly lower HADS than the OffAD-HighHADS group (Tukey HSD: diff=-2.44, p=0.004) (see Figure 6A). Visual inspection of the motor score data on the three measures, across the timepoints separated by these three groups again demonstrates the negative relationship between antidepressant prescription and recovery even relative to the OffAD-HighHADS (see Figures 6B-D).

### No evidence of a relationship between HADS score admission scores or improvement

There was not sufficient evidence to reject the null hypothesis of no relationship between HADS and admission scores (p=0.170, 6/96 models significant, mean β= -0.003, range - 0.004 to -0.001) or improvement (p>0.999, 0/288 models significant, mean β= -0.001, range -0.004 to 0.001).

## Discussion

This retrospective study examined whether patients prescribed different classes of common, CNS-acting, drugs (GABA agonists, sodium or calcium channel blocking antiepileptics, or antidepressants) responded differently to an intensive, high-dose upper-limb rehabilitation programme. To test this robustly, SCA was used, where all sensible variations of models examining a certain hypothesis were run, and the sum of results across all models was interpreted. Using this method patients prescribed GABA agonists were found to have worse upper-limb scores on admission to the programme but did not differ in terms of their improvement. This was in contrast to patients prescribed antidepressants, who did not differ on admission scores but had significantly poorer upper-limb improvement. There was no difference in admission or improvement scores in patients on antiepileptics.

### Patients on GABA agonists had worse admission scores but did not differ in programme-related improvements in function

Across all three upper-limb measures, patients on GABA agonists had significantly worse admission scores, around a 6-10% reduction relative to those not prescribed the drug. Despite the large regression coefficient size, this difference is somewhat difficult to interpret. The drugs in the GABA agonist category are prescribed for diverse problems, for example baclofen (prescribed to 84% of the GABA agonist group) for spasticity or benzodiazepines (18% of GABA agonist group) for anxiety, insomnia and seizures. Clearly any differences in admission scores could be attributed either to the underlying co-morbidity for which the drug is prescribed, the effects of drug itself, or an association between the co-morbidity and increased stroke severity. While there were some control measures recorded at admission, e.g. HADS and NFI scores, there were not any measures of spasticity or sleep quality which might be relevant for assessing differences between those on and off GABA agonists.

Perhaps a more pertinent finding for clinical practice is the lack of significant difference in programme-related improvements in upper-limb function between patients on and off GABA agonists. Several studies have previously reported a correlational link between high GABA concentration (27), or receptor activity (28,29), and worse functional outcomes from rehabilitation post-stroke. Furthermore, a single dose of the GABA_B_ agonist baclofen impairs aspects of motor learning in healthy humans (30); and GABA antagonists can improve post-stroke motor recovery in rats (13,14). Given these findings, and another early retrospective study finding a negative impact of benzodiazepine prescription on motor function recovery (16, though see 31), caution has previously been advised in the prescription of GABA agonists, particularly benzodiazepines, post-stroke (31).

Yet in this data set, patients who were taking GABA agonists did not differ in degree of programme-induced improvements even despite co-morbidities which could additionally hamper potential for improvement from the programme. The result reported here should not, however, be taken as evidence that these drugs do not have any detrimental effects on motor rehabilitation-patients were sometimes advised to take these medications at night, or only as needed, likely minimising their potential to interact with rehabilitation. Rather, this result should be interpreted as the absence of difference in programme-induced improvements for patients with typical GABA agonist prescriptions. It could also be argued that the symptoms which these drugs seek to treat, e.g. spasticity or insomnia, may themselves worsen rehabilitative potential to a greater degree if left unresolved (32). Furthermore, we cannot exclude that our lack of effect is due to low power, and so further large-scale studies are needed.

### Patients on sodium and calcium channel blocking antiepileptics did not significantly differ on admission scores or motor improvements on the QSUL programme

Stroke is the cause of 10% of all epilepsy cases (33) and so a great deal of stroke patients, 29% in this data-set, are prescribed antiepileptics targeting sodium and calcium channels. Here we found that there were no significant differences in admission motor scores for patients prescribed antiepileptics versus those who were not. Comparing improvements on the QSUL programme between the groups also resulted in a non-significant difference, however there was a trend towards a decrease in improvements for patients on antiepileptics. Closer examination of this finding shows that it was driven only by poorer improvements on one measure, the ARAT, with very little effect on the CAHAI or FM, suggesting that this was not a robust effect across motor measures.

Though classic antiepileptic treatments, such as phenytoin or phenobarbital, have been suggested to be detrimental to motor recovery in retrospective studies (16), there is little evidence for any influence of modern antiepileptic drugs on patient outcomes (34). In fact some animal studies have even found neuroprotective benefits of Na channel blockers (35). The results presented here align with a lack of significant effect of this class of drugs on rehabilitation-induced motor improvements when prescribed appropriately.

### Patients prescribed antidepressants do significantly worse on the QSUL programme

Post-stroke depression is a frequent complication of stroke (36,37), most commonly treated by antidepressant prescription. Here we found that there were no significant differences in admission scores between patients with and without antidepressant prescriptions. However, when examining the programme-induced improvements in motor scores, patients on antidepressants did worse than those off the drugs. Significant regression coefficients were evenly distributed across different motor measures, whether examining outcome given baseline or recovery, and whether subjective mood information (i.e. HADS and NFI scores) was included in the model or not.

Poorer motor improvements in patients on antidepressants could be driven by effects of the drugs themselves, of the underlying depression, or a combination of the two. Patients with antidepressant prescription had higher HADS scores, i.e. had more symptoms of depression and anxiety, than those without. However, the persistence of the difference between patients across antidepressant prescription while controlling for HADS, the non-significant relationship between HADS and improvement, and the observation that patients on antidepressants do worse than patients with higher HADS scores but off antidepressants, indicates that there is some relationship specific to this ‘on antidepressants’ category.

This result lies somewhat in contrast to the literature on the effect of SSRIs for post-stroke motor recovery. Inspired by the results of animal (38) and smaller human studies (8–11), one medium sized placebo-controlled trial found that 3 months of 20mg fluoxetine daily, alongside physiotherapy, improved motor outcomes in chronic stroke patients (12), and a similar pattern of positive results has also been found for drugs influencing the noradrenergic system (39). More recent studies without additional universal concurrent physiotherapy have however, reported null results (40–42), leading some to suggest that SSRIs are creating a brain environment conducive for plasticity which can then be exploited by concurrent rehabilitative training (17,43).

Here antidepressants (the vast majority of which were SSRIs, ∼80%) were paired with rehabilitation, and so might be predicted to boost recovery. Some speculative reasons could be proposed for this divergence in findings: it may be that a beneficial effect of SSRIs does not persist in conjunction with depressive symptoms; or it could be that the antidepressant prescription is a better measure of trait depression across the 6 month duration of the follow-up than the one-time HADS score at admission, and the negative impact of these depressive symptoms may outweigh any positive impact of the drug. Additionally, the patients in QSUL programme tended to be several months post stroke and were receiving intensive rehabilitation, whereas randomised controlled trials assessed the influence of SSRIs on acute patient recovery, in the days to weeks after stroke, with (at most) only standard in-patient physiotherapy (12). Further research is needed to identify a mechanistic explanation for the negative relationship, but there is still value in the observation that patients with antidepressant prescriptions tend to do worse on intensive rehabilitation programmes. Identifying those patients who may respond less well to the treatment is the first step in developing methods to improve interventions for these patients.

## Conclusions

This retrospective study investigated the relationships between prescriptions of three classes of commonly used, CNS-acting, drugs and upper-limb improvements of 277 patients during the 3-week intensive QSUL programme. Patients who were prescribed GABA agonist drugs tended to have worse upper-limb scores at admission, but there was no evidence of differences in response to the programme. This indicates that, when appropriately prescribed, patients with GABA agonist prescription did not perform significantly differently on this upper-limb rehabilitation programme. This was in contrast to patients with antidepressant prescriptions where no evidence was found for significantly different upper-limb scores at admission, but these patients showed poorer improvement on the programme that could not be explained by the HADS measure of depression and anxiety. If these patients can be identified prior to admission, then differences in their needs on such programmes may be better identified. There was no evidence of significant differences in patients with or without antiepileptic drug prescriptions on either admission to, or improvement on, the programme. Further research is needed to understand these relationships in more detail and to examine whether the results generalise to other study populations, less intensive upper-limb interventions, and larger-scale samples.

## Data Availability

Data are available for credible researchers upon request to Dr Ward. Code available at https://github.com/ainsliej/SCA-QSUL_Drugs

## Acknowledgements

A Johnstone is funded by a project grant from the Dunhill Medical Trust. Thanks to all the physiotherapists and occupational therapists at The National Hospital for Neurology and Neurosurgery, Queen Square, who have treated patients on this programme. Thanks to UCLH Charities, Friends of UCLH and The National Brain Appeal for funding to purchase equipment used in this programme.

